## Appendix for "Early intervention is the key to success in COVID-19 control"

### Mathematical model specification

This Appendix contains a mathematical specification of the stochastic continuous-time branching process used to model the spread of COVID-19. Model parameters and distributions are given in Table S1.

#### Branching process model

Infected individuals are grouped into two categories: (i) those who show clinical symptoms at some point during their infection; and (ii) those who are subclinical throughout their infection. Each new infection is randomly assigned as subclinical with probability $p_{\text{sub}} = 0.33$ and clinical with probability $1 - p_{\text{sub}}$, independent of who infected them. Once assigned as clinical or subclinical, individuals remain in this category for the duration of their infectious period.

The average reproduction number $R_{\text{sub}}$ of subclinical individuals was assumed to be 50% of the average reproduction number $R_{\text{clin}}$ of clinical individuals. This reflects lower infectiousness of subclinical cases (Davies et al., 2020). Individual heterogeneity in transmission rates was included by setting the individual reproduction number $R_i$ for individual $i$ to be $R_i = R_{\text{clin}} Y_i$ for clinical cases and $R_i = R_{\text{sub}} Y_i$ for subclinical infections, where $Y_i$ is a gamma distribution with mean 1 and variance $1/k$. In the absence of case isolation measures (see below), each infected individual $i$ causes a randomly generated number $N_i \sim \text{Poisson}(R_i)$ of new infections. This is equivalent to the superspreading model of Lloyd-Smith et al. (2005) with dispersion parameter $k$.

The time between an individual becoming infected and infecting another individual, the generation time $T_G$, follows a Weibull distribution with mean 5.0 days and standard deviation 1.9 days (Ferretti et al., 2020). The infection times of all $N_i$ secondary infections from individual $i$ are independent, identically distributed random variables from this distribution. The model does not explicitly include a latent period or pre-symptomatic infectious period. However, the shape of the Weibull generation time distribution captures these phases, giving a low probability of a short generation time between infections and with 90% of transmission occurring between 2.0 days and 8.4 days after infection.

Individuals who have recovered from the virus are assumed to have immunity for the duration of the period simulated and cannot be re-infected. This means that the proportion of the population that is susceptible at time $t$ is $1 - N(t)/N_{\text{pop}}$, where $N(t)$ is the cumulative number of infections at time $t$ and $N_{\text{pop}}$ is the total population size.

Clinical cases have a probability $p_{\text{det}}$ of being tested and isolated. The time $T_{\text{iso}}$ between infection and isolation is modelled as the sum of two independent random variables $T_1$ and $T_2$. $T_1$ represents the incubation period (time from exposure to onset of symptoms) and has a gamma distribution with mean 5.5 days and standard deviation 2.3 days (Lauer et al., 2020). $T_2$ represents the time from symptom onset to isolation and is modelled as an exponential distribution with mean 2.2 days, based on New Zealand case data. Following isolation, transmission is reduced to a proportion $c_{\text{iso}} < 1$ (Davies et al., 2020) of the value it would be without isolation. There is a further delay from isolation until a positive test result is returned, which is assumed to be exponentially distributed with mean 3.48 days. We assume that infected individuals always return a positive result when tested (i.e. we do not model false negatives). Subclinical infections do not get tested or isolated and are not reported as confirmed cases (i.e. they remain undetected). This models a symptom-based testing regime where asymptomatic individuals are not routinely tested, which was largely the case in New Zealand's March-May outbreak.

Each clinical case is assumed to have a fixed probability of resulting in fatality. We simulate this using a Poisson approximation such that a randomly generated number of deaths, $M \sim \text{Poisson}(N_{\text{clin}}\text{IFR}/(1-p_{\text{sub}}))$, occur out of the total number of clinical cases, $N_{\text{clin}}$, where IFR is the infection fatality rate. We use an IFR of 0.88%, obtained by fitting age-specific COVID-19 IFR estimates from international studies (Verity et al, 2020) to the age distribution of the New Zealand population from 2018 Census data (Statistics New Zealand, 2020).

For computational convenience, the continuous-time model defined above is simulated using a time step of $\delta t = 1$ day. At each time step, infectious individual $i$ produces a Poisson distributed number of secondary infections with mean

$$\lambda_i(t) = C(t)\left(1 - \frac{N(t)}{N_{\text{pop}}}\right) R_i F\left(t - T_{I,i} - T_{\text{iso},i}\right) \int_t^{t+\delta t} W\left(\tau - T_{I,i}\right) d\tau, \tag{1}$$

where $T_{I,i}$ is time individual $i$ became infected, $T_{\text{iso},i}$ is the time between infection and isolation of individual $i$, $C(t)$ is the relative population-wide transmission rate at time $t$ (with higher Alert Levels corresponding to smaller values of $C(t)$), and $F(t)$ is a function describing the reduction in transmission due to isolation defined by:

$$F(s) = \begin{cases} c_{\text{iso}}, & s \geq 0 \text{ and } D_i = 1 \\ 1, & \text{otherwise} \end{cases}, \tag{2}$$

where $D_i \sim \text{Bernoulli}(p_{\text{det}})$ for clinical cases and $D_i = 0$ for subclinical cases is a random indicator variable that equals 1 if case $i$ is tested. All individuals are assumed to be no longer infectious 30 days after being infected. This is an upper limit for computational convenience; in practice, individuals have very low infectiousness after about 12 days because of the shape of the generation time distribution.

### Seed cases

The model was initialised with seed cases representing arrival of infected individuals from overseas. For the factual scenario (Scenario 0), the number and timing of these seed cases was chosen to replicate actual imported cases. Data was obtained from the Ministry of Health on confirmed and probable cases of COVID-19 in New Zealand. For each case, the dataset contained the following fields: whether there was a known recent international travel history and, if so, date of arrival to New Zealand; date of symptom onset (from patient recall, where available); date of isolation; date of reporting.

Model simulations were seeded with the $N_{\text{intl}}$ cases with a known recent history of international travel, using their actual dates of arrival, symptom onset, isolation and reporting. For these cases, the infection date was estimated backwards from the date of symptom onset (using the incubation period distribution shown in Table S1). For cases without an onset date, the infection date was instead backdated from the arrival date. Cases without an isolation date that arrived prior to 9 April 2020 were assumed to remain unisolated (i.e. $F(s) = 1$) for the whole infectious period. Secondary infections that occurred before arrival in New Zealand were ignored. Cases that were recorded as having a recent international travel history but missing an arrival date were assumed to have arrived at the same time as infection, i.e. they spent their entire infectious period in New Zealand. All international cases that arrived after 9 April 2020 were placed in government-managed isolation and quarantine (MIQ), which we assume is 100% effective at preventing onward transmission by these individuals.

To allow for the fact that the case data only includes clinical cases, an additional number $N_{\text{intl,sub}} \sim \text{Poisson}\left(N_{\text{intl}} p_{\text{sub}}/(1-p_{\text{sub}})\right)$ of subclinical seed cases were added. The infection dates and arrival dates for these subclinical seed cases were approximated by randomly sampling with replacement from the clinical seed cases. We assume that these international subclinical cases are not detected and therefore do not self-isolate, but those arriving after 9 April are placed in MIQ and do not contribute to local transmission.

To simulate scenarios with border restrictions starting earlier than the actual dates, seed cases arriving from overseas between the earlier start date and the actual start date were assumed to be self-isolated on their date of arrival. To simulate a delay of $n$ days to border closure, we delayed the arrival dates (and associated symptom onset, reporting and isolation dates) of international seed cases arriving after the actual border closure date (19 March 2020) by $n$ days. We then allowed for new international cases arriving over this delay period (e.g. additional non-residents that may have chosen to travel had the border remained open for longer) by seeding an additional Poisson-distributed random number of international cases over the $n$ days after 19 March, with an average daily number of seeded cases equal to the actual average daily number of international cases arriving during the week prior to 19 March (33 international cases per day). These additional seeded cases were assumed to self-isolate on arrival and their delays from arrival-to-symptom onset and arrival-to-reporting were randomly sampled with replacement from the corresponding delays in the actual international case data.

For each simulation, the model outputs the number of newly infected cases each day, the number of cases reported each day and the number of fatalities. These outputs were averaged over $M$ independently initialised realisations of the model. We also report the interval range within which 90% of simulation results were contained.

### Effective reproduction number

In the absence of any case-targeted or population-wide control measures (i.e. with $c_{\text{iso}} = 1$ and $C(t) = 1$), the implied basic reproduction number of the model is

$$R_0 = (1 - p_{\text{sub}})\, R_{\text{clin}} + p_{\text{sub}} R_{\text{sub}} \tag{3}$$

For the values of $p_{\text{sub}}$, $R_{\text{clin}}$ and $R_{\text{sub}}$ shown in Table S1, this gives a basic reproduction number of $R_0 = 2.5$.

Population-wide control measures at time $t$ reduces transmissions by a factor of $C(t)$. Case isolation reduces transmission by a factor of $c_{\text{iso}}$ from a proportion $p_{\text{det}}$ of clinical cases after their isolation time. Together, these measures give an effective reproduction number $R_{\text{eff}}$ in a fully susceptible population of

$$R_{\text{eff}} = C(t)\left[(1 - p_{\text{sub}})\left(p_{\text{det}}\left(q + c_{\text{iso}}(1-q)\right) + 1 - p_{\text{det}}\right) R_{\text{clin}} + p_{\text{sub}} R_{\text{sub}}\right] \tag{4}$$

where $q = \Pr(T_G < T_1 + T_2)$ is the proportion of transmission that occurs prior to isolation. Table S1 shows the values of $C(t)$ used to model New Zealand's four COVID-19 Alert Levels during the March-May 2020 outbreak and the corresponding values of $R_{\text{eff}}$ calculated using Eq. (4).

### Model parameters

*Table S1: Parameters used for model simulations and their sources.*

| **Parameter** | **Value** | **Source** |
|---|---|---|
| Distribution of generation times | $Weibull(\text{scale} = 5.67, \text{shape} = 2.83)$ | Ferretti et al (2020) |
| Distribution of exposure to symptom onset (incubation period) (days) | $\mathrm{T}_1 \sim \Gamma(\text{shape} = 5.8, \text{scale} = 0.95)$ | Lauer et al (2020) |
| Distribution of symptom onset to isolation (days) (from data) | $T_2 \sim Exp(\text{mean} = 2.18)$ | Davies et al (2020a) |
| Distribution from isolation to reporting | $\Gamma(\text{shape} = 1, \text{scale} = 6)$ | Fitted to data |
| Reproduction number for clinical infections (no case isolation or control) | $R_{clin} = 3$ | Davies et al (2020a) |
| Reproduction number for subclinical infections | $R_{sub} = 1.5$ | Davies et al (2020a) |
| Relative infectiousness of subclinical cases | $R_{sub}/R_{clin} = 50\%$ | Davies et al (2020a) |
| Proportion of subclinical infections | $p_{sub} = 33\%$ | Davies et al (2020b); Byambasuren et al (2020); Lavezzo et al (2020) |
| Relative infectiousness after self-isolation (home quarantine); and after MIQ | $c_{iso} = 65\%$; $c_{MIQ} = 0\%$ | Davies et al (2020a) |
| Proportion of clinical cases detected and reported | $p_R = 75\%$ | Price et al (2020); and estimated from limited NZ data |
| Individual heterogeneity in transmission rate | $Y_K = 0.5$ (i.e. moderate super-spreading) | Endo et al (2020); Lloyd-Smith et al (2005) |
| Infection fatality rate (IFR) | 0.88% | Age-specific IFR estimates from Verity et al (2020), fitted to NZ's age distribution from 2018 Census data |
| Transmission rate relative to no population-wide control, at AL 1, 2, 3 and 4; and in the period prior to AL4 | *C(t)* = 1, 0.7, 0.4 and 0.147 (corresponding to $R_{eff}$ = 2.37, 1.66, 0.95 and 0.35); and *C(t)* = 0.75 ($R_{eff}$ =1.8) | Flaxman et al (2020); Binny et al (2020a); Binny et al (2020b); and limited NZ data |

### Supplementary Tables and Results

*Table S2: Timing of national border management and lockdowns in different countries (selected to illustrate the diversity of responses). Border restrictions describe 14-day quarantine restrictions in government-managed isolation and quarantine (MIQ) facilities, or at home, or in another country with little or no community transmission, unless otherwise stated. For most countries, earlier border restrictions were in place for travellers from certain high-risk/COVID-19-affected countries. Border closures are restrictions on the entry of specific categories of travellers (e.g non-citizens/residents, or potentially all arrivals). Lockdowns refer to physical distancing and movement restrictions which may extend to stay at home orders. Dates are given for the first occasion that measures were implemented along with Days after 1st reported case; Total cases refers to the number of cases at the time the measure was introduced. Note that some countries have since lifted and/or reinstated measures.*

| | **Border restrictions** | | | **Border closure** | | | **Lockdown** | | | **100 days after 1st case** | |
|---|---|---|---|---|---|---|---|---|---|---|---|
| **Country/State /Province** | **Date** | **Days after 1st case** | **Total cases** | **Date** | **Days after 1st case** | **Total cases** | **Date** | **Days after 1st case** | **Total cases** | **Cumulative cases** | **Cumulative deaths** |
| Samoa | 15 Mar | No cases | 0 | 20 Mar | No cases | 0 | 26 Mar | No cases | 0 | 0 | 0 |
| Vanuatu | 20 Mar | No cases | 0 | 20 Mar | No cases | 0 | 26 Mar[1,2] | No cases | 0 | 0 | 0 |
| Tonga | 17 Mar | No cases | 0 | 23 Mar | No cases | 0 | 29 Mar | No cases | 0 | 0 | 0 |
| Solomon Islands | 22 Mar | No cases | 0 | 22 Mar | No cases | 0 | None | No cases | 0 | 0 | 0 |
| Fiji | 20 Mar | 1 | 1 | 25 Mar | 6 | 5 | 20 Mar[1] | 1 | 1 | 18 | 0 |
| **New Zealand** | **15 Mar (Self-isol.); 9 Apr (MIQ)** | **18; 43** | **11; 1283** | **19 Mar** | **22** | **53** | **25 Mar** | **28** | **315** | **1504** | **22** |
| Vietnam | 21 Mar | 58 | 88 | 22 Mar | 59 | 95 | 1 Apr | 69 | 212 | 270 | 0 |
| Taiwan | 19 Mar | 58 | 108 | 19 Mar | 58 | 108 | None | - | - | 429 | 6 |
| Iceland | 18 Mar | 19 | 247 | 20 Mar[3,4] | 21 | 330 | None[1,5] | - | - | 1806 | 10 |
| Australia | 16 Mar | 51 | 298 | 20 Mar | 55 | 709 | 23 Mar[6] | 58 | 1709 | 6801 | 95 |

| | | | | | | | | | | | |
|---|---|---|---|---|---|---|---|---|---|---|---|
| Ontario, Canada | National: 25 Mar | 60 | 688 | National: 18 Mar | 53 | 221 | 24 Mar | 59 | 588 | 19,097 | 1446 |
| South Africa | 26 Mar[7] | 21 | 709 | 26 Mar | 21 | 709 | 26 Mar | 21 | 709 | 61,927 | 1354 |
| Japan | 3 Apr | 78 | 2617 | 1 Apr | 76 | 1953 | None[2] | - | - | 12,892 | 551 |
| Hubei, China | National: 28 Mar | 132 | 67,801 | Provincial: 23 Jan; National: 28 Mar | 67; 132 | 444; 67,801 | 23 Jan | 67 | 444 | 64,786 | 2563 |
| Italy | 17 Mar | 46 | 27,980 | 17 Mar[3,4] | 46 | 27,980 | 10 Mar | 39 | 9172 | 218,268 | 30,395 |
| Germany | 10 Apr | 74 | 113,525 | 18 Mar | 51 | 7156 | 23 Mar | 56 | 24,774 | 164,897 | 6996 |
| France | 11 May | 108 | 139,063 | 17 Mar | 53 | 6633 | 17 Mar | 53 | 6633 | 130,979 | 24,760 |
| UK | 8 Jun | 131 | 264,039 | None | - | - | 23 Mar | 54 | 8934 | 199,404 | 30,321 |
| New York, USA | 24 Jun[7] | 115 | 389,666 | None[8] | - | - | 22 Mar | 21 | 15,800 | 379,482 | 30,458 |
| Sweden | None | - | - | 19 Mar | 44 | 1410 | None | - | - | 28,753 | 3745 |
| Brazil | None | - | - | 30 Mar | 33 | 4256 | None[1] | - | - | 614,932 | 34,021 |

[1]*Local lockdown(s) only.*

[2]*Nationwide state of emergency declared ("soft" lockdown).*

[3]*Weak restrictions: travel permitted for work and health reasons.*

[4]*Border closed to all non-EU/EEA citizens.*

[5]*Highly intensive contact tracing and testing regime.*

[6]*Schools remained open in some states, though attendance dropped by up to 50% with many parents choosing to keep children at home.*

[7]*Restrictions apply only to travellers arriving from high-risk/COVID-19-affected countries/states.*

[8]*USA-Canada and USA-Mexico land borders closed to non-essential travel on 21 March and foreign nationals from certain COVID-19-affected countries not permitted to enter.*

*Table S3: Sensitivity tests for alternative values of relative transmission rate C(t)=0.295, 0.4 and 0.4635 (corresponding to $R_{eff}$=0.70, 0.95 and 1.10 respectively) in AL3. Key measures from scenarios of early or delayed implementation of policy interventions: the maximum number of daily new cases, date on which the peak occurs, the number of daily new cases at the end of the Alert Level 4 period, the cumulative number of cases and the total number of deaths at the end of the simulated 7 week period of Alert Level 4/3 restrictions. For each measure, except P(elim), the mean value from 5000 simulations is reported alongside the interval range, in parentheses, in which 90% of simulations results are contained.*

| **Scenario** | **$R_{eff}$ in AL3** | **Max. new daily cases** | **Date of peak** | **New daily cases at end of AL4** | **Cumulative cases** | **Total deaths** | ***P(elim)*, 5 weeks after end of AL3** |
|---|---|---|---|---|---|---|---|
| **Actual** | **Unknown** | **84** | **25-Mar** | **3** | **1503** | **22** | **-** |
| 0 | 0.70 | 80 [67, 98] | 31-Mar [26-Mar, 02-Apr] | 4 [1, 8] | 1442 [1210, 1785] | 23 [14, 33] | 0.79 |
| | 0.95 | 80 [67, 99] | 31-Mar [26-Mar, 02-Apr] | 4 [1, 8] | 1448 [1208, 1796] | 23 [14, 33] | 0.66 |
| | 1.10 | 80 [67, 99] | 31-Mar [26-Mar, 02-Apr] | 4 [1, 8] | 1445 [1212, 1795] | 23 [14, 33] | 0.60 |
| 1 | 0.70 | 69 [61, 79] | 26-Mar [25-Mar, 26-Mar] | 4 [1, 7] | 954 [837, 1140] | 14 [8, 21] | 0.74 |
| | 0.95 | 69 [61, 79] | 26-Mar [25-Mar, 26-Mar] | 4 [1, 7] | 953 [839, 1132] | 14 [8, 21] | 0.63 |
| | 1.10 | 69 [62, 80] | 27-Mar [25-Mar, 26-Mar] | 4 [1, 7] | 954 [838, 1135] | 14 [8, 21] | 0.55 |
| 2a | 0.7 | 107 [85, 138] | 06-Apr [01-Apr, 09-Apr] | 7 [2, 12] | 2374 [1939, 2987] | 39 [26, 55] | 0.71 |
| | 0.95 | 108 [84, 139] | 06-Apr [01-Apr, 09-Apr] | 7 [3, 12] | 2373 [1918, 2999] | 39 [26, 55] | 0.57 |
| | 1.10 | 108 [84, 139] | 06-Apr [01-Apr, 09-Apr] | 7 [3, 12] | 2376 [1928, 3006] | 39 [26, 54] | 0.48 |
| 2b | 0.70 | 179 [137, 235] | 11-Apr [09-Apr, 14-Apr] | 12 [6, 19] | 3998 [3150, 5162] | 68 [48, 92] | 0.55 |

| | | | | | | | |
|---|---|---|---|---|---|---|---|
| | 0.95 | 179 [137, 233] | 11-Apr [09-Apr, 14-Apr] | 12 [6, 19] | 3988 [3161, 5115] | 67 [48, 91] | 0.38 |
| | 1.10 | 179 [137, 235] | 11-Apr [09-Apr, 14-Apr] | 12 [6, 19] | 3992 [3146, 5176] | 67 [48, 92] | 0.33 |
| 2c | 0.70 | 503 [378, 663] | 21-Apr [20-Apr, 23-Apr] | 34 [22, 49] | 11514 [8847, 15038] | 200 [148, 268] | 0.21 |
| | 0.95 | 503 [382, 661] | 21-Apr [20-Apr, 23-Apr] | 34 [22, 49] | 11534 [8854, 15048] | 200 [147, 266] | 0.07 |
| | 1.10 | 503 [377, 659] | 21-Apr [20-Apr, 23-Apr] | 34 [21, 49] | 11525 [8846, 14986] | 200 [147, 264] | 0.045 |
| 3 | 0.70 | 79 [66, 97] | 31-Mar [26-Mar, 02-Apr] | 4 [1, 8] | 1417 [1194, 1755] | 22 [14, 32] | 0.79 |
| | 0.95 | 79 [67, 97] | 31-Mar [26-Mar, 02-Apr] | 4 [1, 8] | 1422 [1194, 1765] | 22 [14, 32] | 0.66 |
| | 1.10 | 79 [66, 97] | 31-Mar [26-Mar, 02-Apr] | 4 [1, 8] | 1414 [1189, 1735] | 22 [14, 32] | 0.62 |
| 4 | 0.70 | 91 [77, 110] | 01-Apr [01-Apr, 02-Apr] | 5 [1, 8] | 1589 [1353, 1946] | 25 [16, 35] | 0.69 |
| | 0.95 | 91 [77, 110] | 01-Apr [31-Mar, 02-Apr] | 5 [1, 9] | 1594 [1359, 1934] | 25 [16, 35] | 0.55 |
| | 1.10 | 88 [74, 107] | 01-Apr [29-Mar, 02-Apr] | 5 [1, 9] | 1597 [1363, 1945] | 25 [16, 35] | 0.47 |
| 5a | 0.70 | 68 [61, 79] | 26-Mar [25-Mar, 26-Mar] | 4 [1, 7] | 940 [828, 1115] | 14 [8, 21] | 0.76 |
| | 0.95 | 68 [61, 79] | 26-Mar [25-Mar, 26-Mar] | 4 [1, 7] | 941 [826, 1119] | 14 [8, 21] | 0.63 |
| | 1.10 | 68 [61, 79] | 26-Mar [25-Mar, 26-Mar] | 4 [1, 7] | 941 [825, 1118] | 14 [8, 21] | 0.55 |
| 5b | 0.70 | 123 [99, 155] | 05-Apr [01-Apr-, 08-Apr] | 7 [3, 13] | 2545 [2096, 3170] | 41 [28, 57] | 0.68 |
| | 0.95 | 120 [97, 152] | 06-Apr [01-Apr, 08-Apr] | 7 [3, 12] | 2501 [2069, 3121] | 41 [28, 56] | 0.53 |
| | 1.10 | 124 [100, 157] | 06-Apr [02-Apr, 08-Apr] | 7 [3, 13] | 2585 [2124, 3217] | 42 [29, 58] | 0.41 |

*Table S4: Summary of restrictions under New Zealand's COVID-19 four-tier Alert Level system and an elimination strategy. Published on the Covid19.govt.nz website. Essential services (including supermarkets, health services, emergency services, utilities and goods transport) continue to operate at all Levels.*

| **Alert Level** | **Measures** |
|---|---|
| Level 4 - Lockdown | • Stay at home except for essential personal movement<br>• Safe recreational activity allowed in local area.<br>• Travel is severely limited.<br>• All gatherings cancelled and public venues closed.<br>• Businesses closed, except essential services and lifeline utilities.<br>• Educational facilities closed.<br>• Rationing of supplies and requisitioning of facilities possible.<br>• Reprioritisation of healthcare services. |
| Level 3 - Restrict | • Stay at home except for essential personal movement - including going to work, school or for local recreation.<br>• Work from home if possible.<br>• Low risk local recreation activities allowed.<br>• Inter-regional travel highly limited (e.g. allowed for essential workers, with limited exemptions for others).<br>• Public venues closed (e.g. libraries, museums, cinemas, food courts, gyms, pools, playgrounds, markets).<br>• Gatherings of up to 10 people allowed but only for wedding services, funerals and tangihanga, with physical distancing and public health measures maintained.<br>• Two-metre physical distancing outside home. One metre in controlled environments, e.g. schools and workplaces.<br>• People must stay within their immediate household bubble, but can expand this to reconnet with close family/whānau, or bring in caregivers, or support isolated people. This extended bubble should remain exclusive.<br>• Businesses can open premises, but cannot physically interact with customers.<br>• Schools (years 1 to 10) and Early Childhood Education centres can safely open but with limited capacity. Children should learn from home if possible.<br>• Healthcare services use virtual, non-contact consultations where possible.<br>• People at high risk of severe illness (older people or those with pre-existing medical conditions) are encouraged to stay at home and take extra precautions when leaving home. They may choose to work. |
